# Protocol for the development of a tool (INSPECT-SR) to identify problematic randomised controlled trials in systematic reviews of health interventions

**DOI:** 10.1101/2023.09.21.23295626

**Authors:** Jack Wilkinson, Calvin Heal, George A. Antoniou, Ella Flemyng, Zarko Alfirevic, Alison Avenell, Virginia Barbour, Nicholas J L Brown, John Carlisle, Mike Clarke, Patrick Dicker, Jo Dumville, Andrew Grey, Steph Grohmann, Lyle C Gurrin, Jill A Hayden, James Heathers, Kylie E Hunter, Toby Lasserson, Emily Lam, Sarah Lensen, Tianjing Li, Wentao Li, Elizabeth Loder, Andreas Lundh, Gideon Meyerowitz-Katz, Ben W Mol, Neil E O’Connell, Lisa Parker, Barbara K. Redman, Anna Lene Seidler, Kyle A Sheldrick, Emma Sydenham, David J Torgerson, Madelon van Wely, Rui Wang, Lisa Bero, Jamie J Kirkham

**Author notes:** Joint senior authorship (Lisa Bero, Jamie J Kirkham).

## Abstract

**Introduction:** Randomised controlled trials (RCTs) inform healthcare decisions. It is now apparent that some published RCTs contain false data and some appear to have been entirely fabricated. Systematic reviews are performed to identify and synthesise all RCTs that have been conducted on a given topic. While it is usual to assess methodological features of the RCTs in the process of undertaking a systematic review, it is not usual to consider whether the RCTs contain false data. Studies containing false data therefore go unnoticed and contribute to systematic review conclusions. The INSPECT-SR project will develop a tool to assess the trustworthiness of RCTs in systematic reviews of healthcare related interventions.

**Methods and analysis:** The INSPECT-SR tool will be developed using expert consensus in combination with empirical evidence, over five stages: 1) a survey of experts to assemble a comprehensive list of checks for detecting problematic RCTs, 2) an evaluation of the feasibility and impact of applying the checks to systematic reviews, 3) a Delphi survey to determine which of the checks are supported by expert consensus, culminating in 4) a consensus meeting to select checks to be included in a draft tool and to determine its format, 5) prospective testing of the draft tool in the production of new health systematic reviews, to allow refinement based on user feedback. We anticipate that the INSPECT-SR tool will help researchers to identify problematic studies, and will help patients by protecting them from the influence of false data on their healthcare.

## Background

Randomised controlled trials (RCTs) are used to assess the benefits and harms of interventions. Systematic reviews of health and care interventions include all RCTs relating to the review question, synthesising the evidence to arrive at an overall conclusion about whether an intervention is effective works and whether it causes harm. It is well-recognised that some RCTs included in systematic reviews of health interventions are unreliable due to methodological limitations. However, relatively little attention has been given to the fact that some published RCTs are untrustworthy not because of methodological limitations, but rather because they contain false data, and may not have taken place at all. This could be due to research misconduct (including fabrication or falsification of data) or critical errors which would not be identified during established assessments of methodological quality.

A recent illustrative example is ivermectin for treatment and prophylaxis of COVID-19. Several systematic reviews evaluating ivermectin for COVID-19 concluded that the drug reduced mortality (1, 2). Subsequently, it became apparent that these systematic reviews had accidentally included RCTs which appear to have been partially or wholly fabricated(3). For example, the spreadsheet purportedly containing the data from one of these trials featured repeating blocks of data(4). Once these RCTs were excluded, the conclusion of a clear benefit of ivermectin was no longer supported(5). The threat posed by RCTs of questionable veracity is not confined to a particular field of medicine or health. For example, studies of this nature have been identified in systematic reviews of Vitamin K for prevention of fractures(6, 7), tranexamic acid for prevention of postpartum haemorrhage(8), and psychological therapies for management of chronic pain(9).

While RCTs are routinely appraised on the basis of both their internal and external validity during the systematic review process, this appraisal is predicated on the assumption that the studies are genuine; the veracity of the studies is not formally assessed. It is now clear that many studies of questionable veracity describe sound methodology, and so are not flagged by critical appraisal frameworks such as Risk of Bias tools. This prompts the question of how we should assess the veracity of RCTs during the systematic review process. The overall aim of the INSPECT-SR (INveStigating ProblEmatic Clinical Trials in Systematic Reviews) project is to develop and evaluate a tool for identifying these *problematic studies* in the context of systematic reviews of RCTs of health interventions. In the following, we give an overview of the project methods.

## Methods/design

The INSPECT-SR tool will be developed using expert consensus in combination with empirical evidence, over five stages (Figure 1): 1) a survey of experts to assemble a comprehensive list of checks for detecting problematic RCTs, 2) an evaluation of the feasibility and impact of applying the checks to RCTs in systematic reviews, 3) a Delphi survey to determine which of the checks are supported by expert consensus, culminating in 4) a consensus meeting to select checks to be included in a draft tool and to determine its format, and finally 5) prospective testing of the draft tool in the production of new health systematic reviews, to allow refinement based on user feedback.

**Figure 1:**
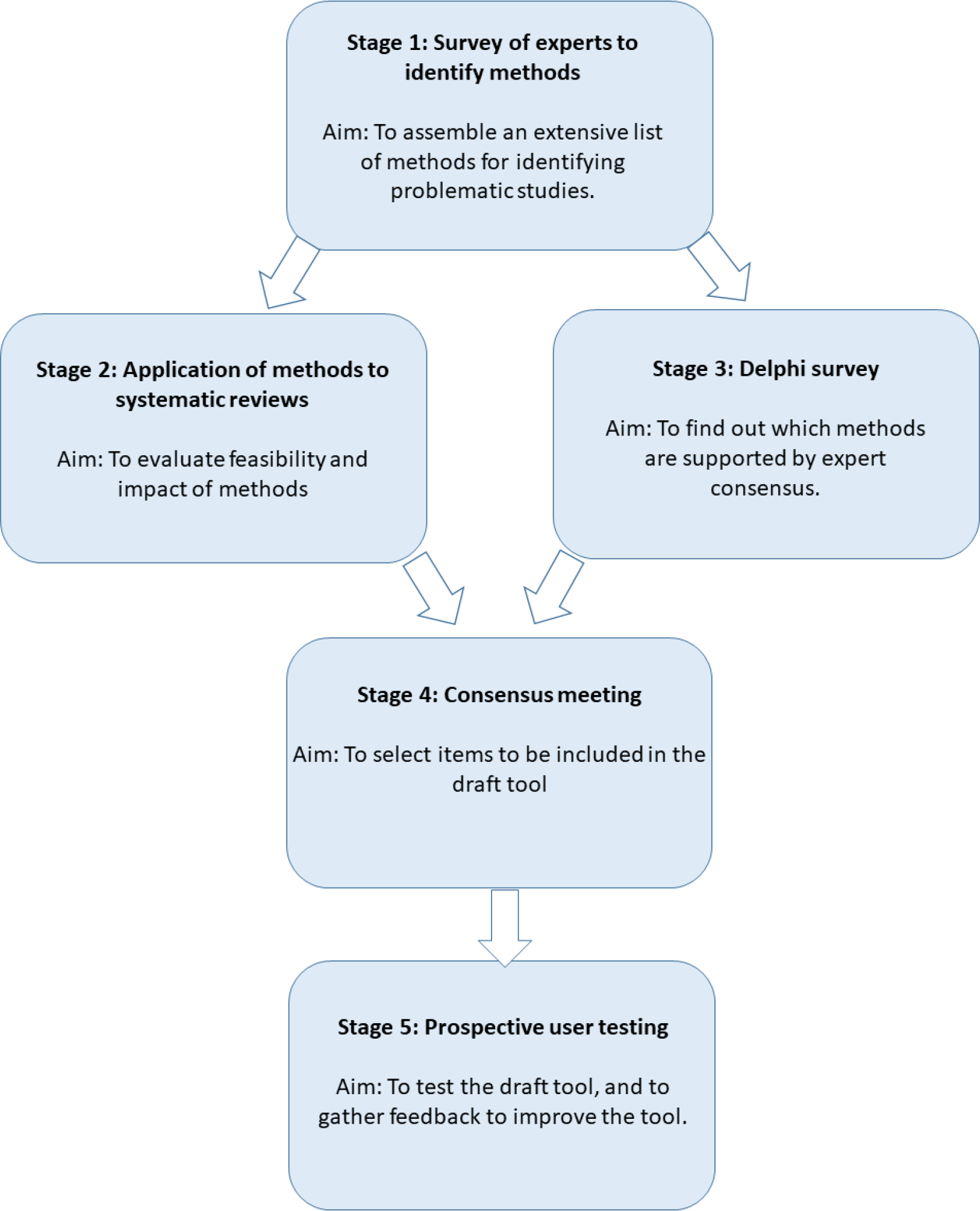
INSPECT-SR development process.

### Working definition of a ‘problematic study’

The Cochrane policy on Managing Potentially Problematic Studies (10, 11) defines a problematic study as “any published or unpublished study where there are serious questions about the trustworthiness of the data or findings, regardless of whether the study has been formally retracted.” We adopt this as a working definition at the outset of the INSPECT-SR project, noting that the project involves the identification of criteria for evaluating ‘trustworthiness’. Criteria under consideration could include statistical checks of data and results, aspects of research governance such as ethical approval, presence of plagiarised content, plausibility of the study conduct, or the track record of the research team. Criteria relating to internal or external validity of results produced by RCTs, such as those included in Risk of Bias(12), Risk of Bias 2(13), or GRADE(14) frameworks, are not within the scope of the INSPECT-SR project. The INSPECT-SR tool will be designed to be used alongside these established critical appraisal frameworks. Assessment of conflicts of interest will be covered by TACIT (Tool for Addressing Conflicts of Interest in Trials)(15) and will not be covered by INSPECT-SR.

### INSPECT-SR working group

The INSPECT-SR working group comprises a steering group, an expert advisory panel, a Delphi panel, and additional collaborators. The steering group includes experts in research integrity, clinical trials methodology, systematic reviews, consensus methodology, and methodological guideline development. They will coordinate the development and evaluation of INSPECT-SR. A larger expert advisory panel has been established to provide advice and to contribute throughout the project. This expert panel has been selected to represent a diverse range of relevant expertise and experience. This includes methodologists, research integrity specialists, public contributors, researchers with experience of investigating potentially problematic studies, experts in systematic reviews, and journal editors. Members of the steering group and expert advisory panel were involved as participants in the Stage 1 survey, and may be eligible to participate in the Stage 3 Delphi survey and consensus meeting. Additional collaborators are involved in Stage 2, and may be eligible to participate in the Stage 3 Delphi survey.

### Stage 1: Survey of experts to assemble an extensive list of checks

#### Overall design

The Stage 1 survey of experts has been completed at the time of writing, and a short protocol for the survey has been posted online (https://osf.io/6pmx5/). We describe the methods briefly here. The aim of Stage 1 was to create an extensive list of checks for identifying problematic research studies, which could be taken forward for evaluation in stages 2 and 3. In Stage 1, we did not restrict our focus to checks applicable to or designed for RCTs specifically. Instead, we sought to identify checks applicable to any research design, so that these could be subsequently evaluated for their applicability to RCTs.

We assembled an initial list of 102 checks that could be used to assess potentially problematic studies. The initial list included checks identified in a recent scoping review(16), a recent qualitative study of experts(17) and additional methods known to the research team (for example, JW undertakes integrity investigations for scientific journals and publishers, and added checks known to him as a result of this work). The list was grouped into several preliminary domains, as shown in Table 1 (adapted from https://osf.io/6pmx5/).

**Table 1:** Frequency of items grouped into preliminary domains included in an online survey of experts (Stage 1).

| Preliminary domain | Frequency of items per domain |
| --- | --- |
| 1. Inspecting results in the paper | 26 |
| 2. Inspecting the research team | 17 |
| 3. Inspecting conduct, governance and transparency | 19 |
| 4. Inspecting text and publication details | 7 |
| 5. Inspecting individual participant data | 33 |
|  | 102 |

We incorporated the list in an online survey in Qualtrics (available at https://osf.io/6pmx5/) to identify checks which had not already been included on the list, and to allow respondents to comment on the checks which were on the list.

The survey asked participants about their experience in assessing potentially problematic studies and to state the country in which they primarily work, before presenting them with the initial list of 102 checks. Each item was presented alongside a free-text box, and participants were advised to comment on any aspect if they wished to do so. At the end of the list, participants were asked whether they were aware of any other checks that had not featured on the list, and were presented with a free text box to describe these.

#### Participants and recruitment

People with expertise or experience in assessing potentially problematic studies, before or after publication, were eligible to participate in the survey. We identified eligible individuals primarily through professional networks, including promotion of the project via conference presentations, and by social media. Members of the steering group and expert advisory panel were invited to participate. We invited eligible individuals by personalised email, and asked whether they could suggest any other potential participants. We attempted to achieve global representation by monitoring the countries in which the respondents worked as responses accumulated, and renewing our efforts to identify and recruit respondents from underrepresented regions. We targeted a minimum sample size of 50, but obtained as many responses as possible.

#### Analysis and next steps

Descriptive analysis of the survey participants (country of work, experience with assessing potentially problematic studies) and responses will be performed. Additional items suggested by respondents, and comments made on existing items, will be summarised. Based on the survey responses further items will be added to the list, and the wording of existing items will be amended, subject to review by the steering group and expert advisory panel members. The updated list will be taken forward to Stages 2 and 3.

Checks categorised in Domain 5 (Inspecting individual participant data, see Table 1) may only be performed when the underlying dataset for an RCT can be obtained. An extension to the INSPECT-SR tool containing Domain 5 checks is in development (working name *INSPECT-IPD*). The development of INSPECT-IPD requires a different approach to the main INSPECT-SR tool (application of checks to a large sample of individual participant datasets, and a distinct Delphi panel). The remainder of this protocol describes the development of the INSPECT-SR tool, which will include checks in Domains 1 to 4 only.

### Stage 2: Retrospective application of the list of items to systematic reviews

#### Overall design

We will apply the full list of checks we have identified to RCTs in a large sample of systematic reviews of interventions published in the Cochrane library, in order to evaluate their feasibility and impact.

#### Review selection

We will use a sample of 50 Cochrane Reviews. This sample size has been selected on a pragmatic basis, to allow a sufficient number of applications of the checks to evaluate feasibility and to characterise the impact on results, while remaining achievable. Stage 2 will be undertaken as a large collaborative enterprise, with steering group members, expert advisory panel members, and additional collaborators who have expressed an interest in participating, each applying the full list of checks to the RCTs in a small number of Cochrane Reviews.

We will endeavour to match assessors to topic areas with which they have familiarity, as this reflects how the final INSPECT-SR tool would be used. We will ask each assessor to state a broad topic area relating to their expertise. We will then identify the most recent Cochrane Review relating to this topic and meeting the eligibility requirements. Where an assessor does not have a particular topic of interest, we will select a topic in order to achieve broad coverage of subjects, and will identify the most recently published Cochrane Review meeting the eligibility requirements. To be eligible, a Review cannot be authored or co-authored by the assessor, out of concern that this could introduce an incentive to overlook problematic features of included studies. Similarly, the Review should not contain RCTs authored or coauthored by the assessor. The Review must also contain at least one meta-analysis containing between one and five RCTs as a feasibility constraint. We also require that the Review has not undergone an assessment to identify potentially problematic studies already, as this may have resulted in removal of problematic trials from the meta-analyses. We acknowledge however that this final criterion may frequently be unclear.

#### Data capture

A bespoke data capture form has been produced. Assessors will extract data for each RCT contained in the first meta-analysis in the Cochrane Review which includes between 1 and 5 RCTs. Assessors will initially record their level of familiarity with the topic of the Review (little or no familiarity, some familiarity, high familiarity), and basic information about each RCT, including a study ID based on the names of the first authors of the Review and of the trial, and years of publication of both, and the year of publication of the RCT. Assessors will then extract data for that RCT from the meta-analysis, including sample size per treatment arm and outcome data per treatment arm (e.g. mean and standard deviation for each treatment arm for continuous outcomes, and frequency of events for binary outcomes). The Risk of Bias assessments for that RCT from the Review will be extracted for each domain, as will the corresponding GRADE assessment for the meta-analysis (if there is one).

Assessors will then attempt to apply items from the list of checks from Stage 1 to the RCT. Assessors will be given the opportunity to apply each check, with the exception of checks which require authors of the RCT to be contacted. For each check, assessors will select a response from the options “not feasible”, “passed”, “possibly fail” or “fail”. A free text box will be available for each check so that the assessor may record the reason for their assessment. Finally, having worked through the list of checks, the assessor will record whether they have concerns about the authenticity of the RCT (with options “no”, “some concerns”, “serious concerns”, or “don’t know”), whether they performed any additional checks not included in the list (and if so, what these checks were and what the outcomes were), as well as being given the opportunity to make any additional comments and to estimate how long it took to perform the assessment.

To assist with applying the checks, each assessor will be provided with a guidance document briefly explaining the rationale for each check and instructions on how to apply them. An Excel workbook will be supplied, which can be used to perform some of the statistical checks.

#### Statistical analysis

We will calculate the frequency of each response option for each check (how often each was considered infeasible, how often each one was failed, possibly failed or passed). We will summarise the overall RCT-level assessments of the assessors after applying the checks (whether or not they had concerns about authenticity). We will evaluate the impact of removing trials flagged by each item, by comparing the data included in the primary meta-analysis before and after the application of the method (e.g. numbers of trials, numbers of events, sample size) as well as the results (changes in pooled estimate, confidence interval width, heterogeneity). We will visualise the clustering of checks, by plotting trial-level assessments for each check in an array. We will consider the relationship between the assessments and the risk of bias (for each domain) in the reviews, to understand the relationship between indicators of problems on the one hand and assessments of evidence quality on the other. This will be undertaken using multinomial regression to assess the association between assessment and risk of bias ratings for each risk of bias domain. GRADE assessments refer to collections of trials rather than individual trials, and so we will use ordinal regression to assess the association between the number of trials in the meta-analysis flagged and the GRADE rating.

### Stage 3: Delphi survey

#### Overall design

A two-round Delphi survey will be conducted to determine which checks are supported by expert consensus.

#### Participants and recruitment

Delphi participants will be identified through professional networks of the steering group and expert advisory panel. We will also invite eligible individuals identified and involved in previous stages of the project. We will recruit individuals representing key stakeholder groups, including: individuals with experience or expertise in assessing problematic studies, journal editors, research integrity specialists, systematic reviewers, clinical trialists, and methodologists. We will categorise participants into two larger groups: 1) individuals with expertise or experience in assessing potentially problematic studies and 2) potential users of a tool for assessing potentially problematic studies, noting that participants may be included in both categories. Individuals will be invited via personalised email describing the Delphi survey in the context of the wider INSPECT-SR development project. We will monitor recruitment across stakeholder groups and geographical location, and will attempt to improve recruitment for groups in which recruitment numbers are low by targeting potential participants in these groups. We consider at least 30 expert participants in each of the two participant groups (experts and potential users) to represent the minimum for a credible Delphi. However, ideally we will aim for a minimum of 100 participants overall.

#### Selection of items

The list of items obtained from Stage 1 will be entered into the Delphi survey. Checks will be categorised and presented in several domains (see Table 1 for the preliminary categorisation scheme, used in Stage 1 but subject to change as the project evolves). We will develop suitable language to clearly describe the checks. The list will be approved by the expert advisory panel before we launch the Delphi survey, including review by public contributors to confirm clarity. We will write an explanation to accompany each item, which participants may review if they are unsure of its meaning.

#### Round 1

We will send participants a personalised email outlining the project, together with a link to the survey, which will be implemented online using suitable software. The survey will include the list of checks. In Round 1, respondents will be asked for basic demographic information, to allow categorisation based on domain(s) of expertise. Respondents will be asked to score each check 1 (lowest score) to 9 (highest score) in two dimensions: usefulness and feasibility. Usefulness will relate to the potential effectiveness of the check for detecting a problematic study. Feasibility will relate to the perceived ease of implementation of each check. Participants will also be given the option to indicate that they do not know whether a check is useful or feasible (because, for example, they are unfamiliar with the approach or lack expertise to comment on a particular check). A free-text box will be provided with each check, so that participants may leave any general comments (such as an explanation for their assessment, or suggestions to modify the wording). Round 1 participants will be invited to suggest additional checks.

#### Round 2

In Round 2, we will add any suggested additional checks to the list (subject to review by the steering group and expert advisory panel), and for each item respondents will be presented with both their own scores (1 to 9) and the distribution of scores from the previous round. Participants who were invited to participate in Round 1 but who did not respond will be invited to the Round 2 survey, and will be presented with the distribution of scores from the previous round only. Participants will be asked to provide a new score in light of this information. The Round 2 survey will include a free-text box for each check so that participants may elaborate on their responses.

#### Analysis

Check-specific scores from Round 2 will be summarised for the overall Delphi panel and by stakeholder group. Any items that meet a consensus criterion, defined as scoring 7 or more by at least 80% of participants overall or in one or more stakeholder groups for usefulness, will be automatically considered during the Stage 4 consensus meeting. Items failing to meet a consensus criterion will be discussed by the steering group and expert advisory panel in light of the Stage 2 application exercise, and will be considered for inclusion in the Stage 4 consensus meetings. Feasibility scores will be summarised for each check, and will be used in Stage 4.

### Stage 4: Consensus meetings

Consensus meetings will be held to finalise the checks to be included in the draft INSPECT-SR tool. We anticipate that multiple meetings will be necessary in order to accommodate international time differences. Meetings may be virtual, in-person, or a combination of both. At these meetings, the results of the Stage 2 application exercise and Stage 3 Delphi survey will be discussed, with the purpose of finalising the items to be included in the draft INSPECT-SR tool. The feasibility assessments from the Stage 2 application exercise and Stage 3 Delphi survey will be considered for all items discussed. Items that are considered useful but challenging to implement may not be incorporated into the main tool, but instead included as an optional or recommended check in the accompanying guidance document. Participants will be invited to reflect the range of key stakeholder groups, as described above. We anticipate that 20 to 30 participants will participate in the consensus meetings, with ten to fifteen participants representing each of the two main participant groups (experts and potential users). In addition to determining the checks to include in the tool, it will be necessary to determine its form and structure, and the recommended process for applying it during the systematic review process. It may be necessary to hold additional meetings focussed on these questions.

### Stage 5: Prospective testing of draft tool

#### Overall design

In collaboration with systematic reviewers, we will prospectively evaluate the draft tool by using it in the production of a cohort of new systematic reviews and systematic review updates. The impact of the draft tool’s Impact on Review conclusions will be assessed in the same way as in Stage 2. We will assess feasibility and usability by implementing surveys regarding experiences of use. Separate surveys will be designed for review authors and, for Cochrane Reviews, editors. These will explore ease of implementation, barriers to use, and suggestions for improvement. In addition to user-level data, we will capture data relating to the individual reviews in which the tool was implemented, as each one represents a potentially informative case study. We will undertake additional qualitative interviews with users during this testing phase, to capture additional feedback.

We will aim to include a variety of topic areas in this testing phase. Stage 5 will culminate in a user workshop, including review editors and review authors involved in testing the tool.

#### User workshop

Findings from the surveys will be fed back to participants as part of a user workshop. The workshop might be virtual, in-person, or a combination of both. Participants will share their experiences of using the tool, and make recommendations for refinement. The discursive format of the meeting is intended to reveal additional information about the experience of users that could not be easily captured via the surveys. We will invite both authors and editors involved in the testing phase to participate. The findings of the testing phase will be used to make final modifications to the tool for usability. We will use the results to produce guidance relating to use of the tool in practice. Alongside Stage 5, as we gather user data, we will produce training materials (to be delivered as workshops and as an online training module) to familiarise systematic review authors and editors with the tool. These will be finalised in light of the findings from the user workshop.

## Conclusion

Systematic reviews of health interventions are considered to represent a very high standard of evidence and frequently inform policy and practice. However, because the veracity of included RCTs is not usually considered, systematic reviews may unintentionally act as a pipeline for false data with the risk that this will influence care. While the need to prevent problematic studies from contributing to systematic reviews is recognised, with several recent laudable efforts to tackle the issue (18-20), there is currently limited agreement on how this should be done. The INSPECT-SR project will develop a tool for evaluating the trustworthiness studies, backed by empirical evidence and expert consensus. We anticipate the draft tool will be available early 2024, and the final tool will be available late 2024.

## Data Availability

This is a research protocol. Datasets arising from the project will be made freely available.

## Ethical approval

The University of Manchester ethics decision tool was used, and this returned the result that ethical approval was not required for this project (30^th^ September 2022), which incorporates secondary research and surveys of professionals about subjects relating to their expertise.

